# The Risk of Indoor Sports and Culture Events for the Transmission of COVID-19 (Restart-19)

**DOI:** 10.1101/2020.10.28.20221580

**Authors:** Stefan Moritz, Cornelia Gottschick, Johannes Horn, Mario Popp, Susan Langer, Bianca Klee, Oliver Purschke, Michael Gekle, Angelika Ihling, Rafael Mikolajczyk

## Abstract

Nearly all mass gathering events (MGEs) worldwide have been banned since the outbreak of SARS-CoV-2 as they are supposed to pose a considerable risk for transmission of COVID-19. We investigated transmission risk of SARS-CoV-2 by droplets and aerosols during an experimental indoor MGE (using N95 masks and contact tracing devices) and conducted a simulation study to estimate the resulting burden of disease under conditions of controlled epidemics. The number of exposed contacts was <10 for scenarios with hygiene concept and good ventilation, but substantially higher otherwise. Of subsequent cases, 0%-23% were attributable to MGEs. Overall, the expected additional effect of indoor MGEs on burden of infections is low if hygiene concepts are applied and adequate ventilation exists.

**One Sentence Summary:** Seated indoor events, when conducted under hygiene precautions and with adequate ventilation, have small effects on the spread of COVID-19.

## Introduction

In most countries, the ban of mass gathering events (MGE) was one of the first countermeasures undertaken by governments (*1*). In Germany, early in March 2020, the government issued a general ban of MGEs with more than 1000 people (*2*). With 129 billion Euro turnover in 2019, the event sector is the sixth largest economic sector in Germany and up to 1.5 million people depend on this industry (*3*). Insolvencies in this field will not only have an economic impact, but may also result in the loss of creative skills, training infrastructures and upcoming young athletes. The impact of this loss is not restricted to individuals, but affects an important dimension of the society as a whole. Some observations suggest that the type of an event determines its potential for spreading infectious diseases, for example that the risk of an outbreak related to religious events is higher than for sports events or concerts (*4–6*).

Severe acute respiratory syndrome coronavirus 2 (SARS-CoV-2), causing COVID-19, can be transmitted via droplets, aerosols or through contaminated surfaces (*7–11*). While the debate on relevance of various transmission routes for the spread of COVID-19 is still ongoing (*12–14*), overall, physical proximity and hygiene determine transmission. For assessing droplets based transmission, reported or measured contacts can be used; for studying aerosols, type of activity (and resulting inhalation of the sources and recipient), air flows and room ventilation play an additional role (*15–17*).

We conducted an experimental pop concert with three different hygiene concepts and measured the contacts of each spectator during the event using contact tracing devices (CTD). Furthermore, we developed a computer model of the indoor arena and simulated the aerosol distribution and resulting exposure. Finally, we estimated the additional effect of indoor MGEs on the overall burden of infections using an individual based model. We have considered various aspects of this epidemic including the effects of different hygiene measures, wearing of masks, event sizes, ventilation systems and different baseline incidences.

## Results

### Experiment

On August, 22^nd^ 2020, 1212 individuals participated at a pop concert in the Leipzig Arena (Table S1). All participants and involved staff provided negative results for SARS-CoV-2 tests conducted within 48 hours preceding the event. All wore N95 masks during the event.

Three different scenarios were investigated: 1) No restrictions (the pre-pandemic setting), 2) moderate restrictions (checkerboard pattern seating, twice as many entrances as in 1), 3) strong restrictions (pairwise seating with 1.5 m interspace to the next pair, four times as many entrances).

Each scenario consisted of entry (60 minutes), 1^st^ half (20 minutes, upscaled to 45), half time (20 minutes) including simulated catering, 2^nd^ half (20 minutes, upscaled to 45), and exit (15 minutes). Contacts within a radius of 1.5 m were measured with a CTD. When all contacts (>10s) were considered, the number of contacts was high; when critical contacts with a duration of more than 15 minutes were counted (based on standard definition for contact tracing (*18*)) the numbers decreased below 10 (Fig 1a, Table S2).

**Fig. 1:**
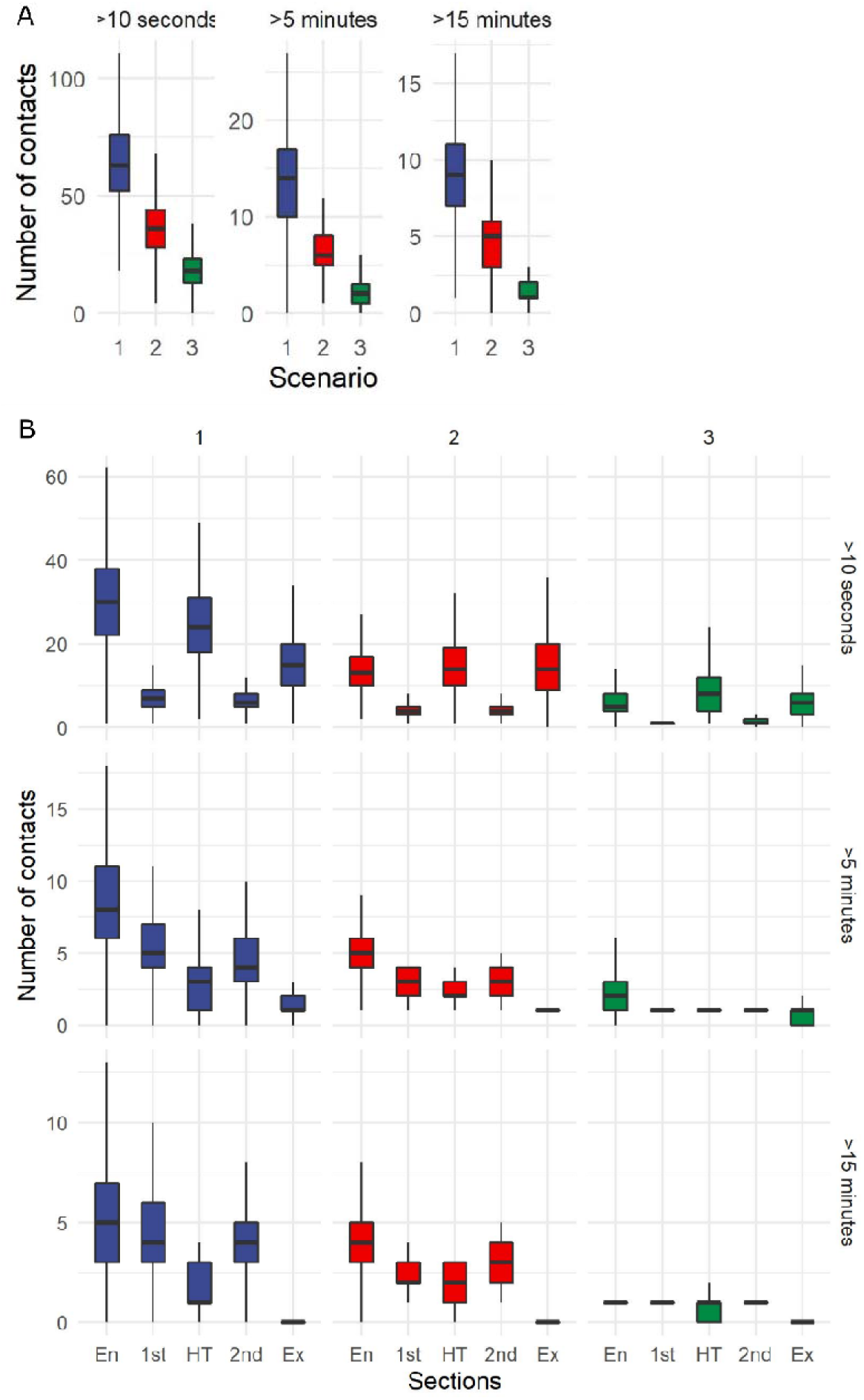
Number of contacts in scenarios 1-3 and by duration (>10 s, >5 min, >15 min). A) Overall, B) during the different sections: Entry (En), 1^st^ half (1^st^), half time (HT), 2^nd^ half (2^nd^), exit (Ex).

High numbers of contacts were observed during entry and half time, but only few lasted more than 15 minutes (Fig. 1b). Hygiene concepts in scenarios 2 and 3 resulted in strong reduction of contacts. Although the exit phase increased mixing of individuals, no contacts over 15 minutes were recorded. In contrast, nearly all contacts during halves lasted longer than 15 min. In scenario 1, new contacts accumulated during the whole event, while in scenarios 2 and 3 most contacts occurred during the entry phase without major further increases (Fig. 2).

**Fig. 2:**
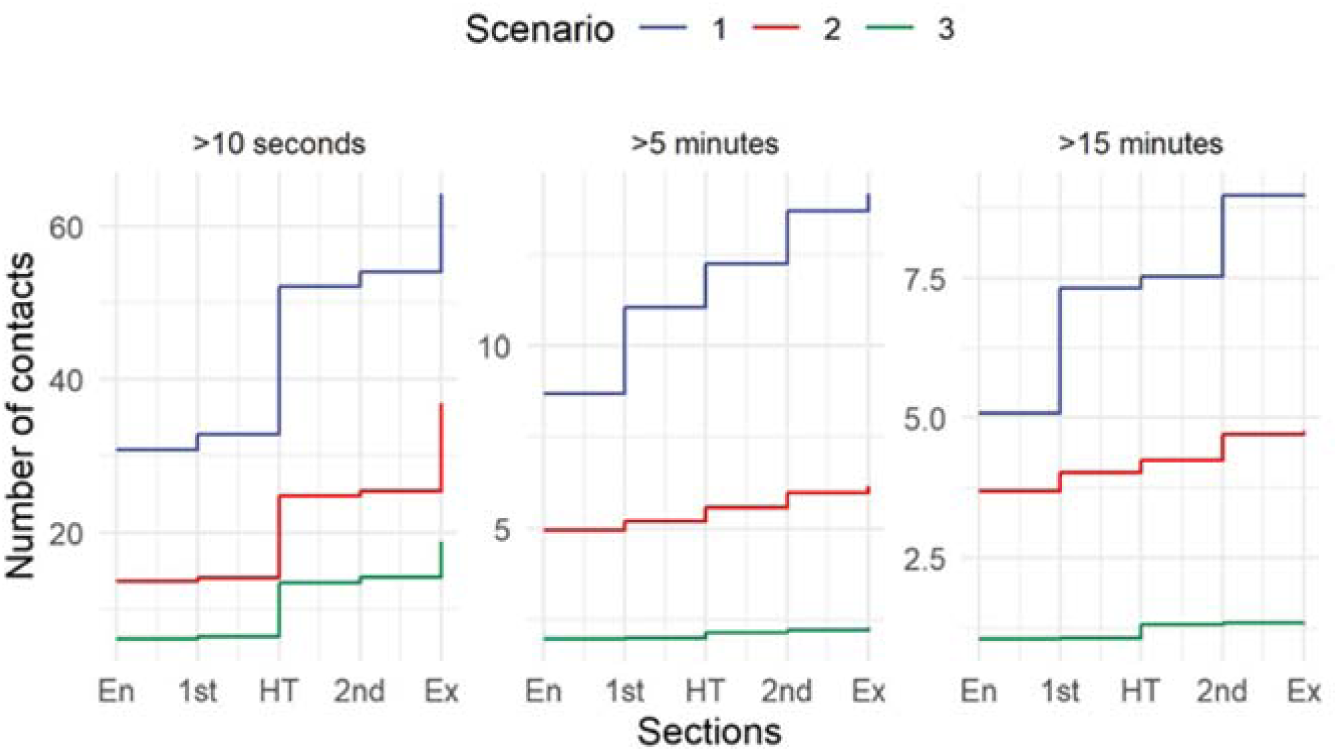
Cumulative number of new contacts across the different sections: Entry (En), 1^st^ half (1^st^), half time (HT), 2^nd^ half (2^nd^), exit (Ex).

### Aerosol exposure

In addition to contacts in physical proximity measured by CTD, we investigated the number of persons exposed to aerosols with a computational fluid dynamics (CFD) simulation. In the simulated current ventilation version (VV) of the arena (VV 1), particle tracking revealed that jet nozzles placed in the arena produce large air rollers on the laterals grandstands. The ejected air streams down from the corner of the roof just above the highest rows, runs parallel to the lateral grandstand to the inner floor of the arena where it rises up to the roof top and the air flow cycle is renewed (Movie S1). Additionally, jet nozzles substantially increase the airflow and thus reduce density of aerosols. We compared the current airflow with a second variant of the ventilation system in which the air was suctioned at the roof top and the jet nozzles were shut down to reduce air rollers in the arena (VV 2). Here, the intended increased vertical airflow from the bottom to the roof (layer ventilation) was not achieved. The lateral inflow of the air resulted in rollers over the lateral grandstands, which were, due to a lower air exchange rate (1.46 vs. 0.85 /h), smaller and slower than in VV 1 (Movie S2). Despite the presence of larger air rollers, the maximum number of exposed people per infectious person was 10 with jet nozzles and higher airflow in VV 1, and 108 in VV 2 for scenario 1. Figure S1 shows an example of the east grandstand with both variants. Furthermore, hygiene concepts of scenario 2 and 3 reduced the number of exposed visitors in both ventilation variants (Table 1).

**Table 1.** Mean number of aerosol exposed individuals of each infected individual for both ventilation versions (VV). Numbers are shown for the three main stands of the arena (stalls, west grandstand, east grandstand) and all stands. SD = standard deviation.

| Stand | Mean number of exposed per each infected individual (±SD) in VV 1 | | | Mean number of exposed per infected individuals (± SD) in VV 2 | | |
| --- | --- | --- | --- | --- | --- | --- |
|  | Scenario 1 | Scenario 2 | Scenario 3 | Scenario 1 | Scenario 2 | Scenario 3 |
| **Stalls** | 3.4 (±2.9) | 1.6 (±1.3) | 0.6 (±0.9) | 12.1 (±6.1) | 5.3 (±3.0) | 2.9 (±2.3) |
| **West grandstand** | 4.8 (±3.5) | 2.6 (±1.8) | 1.1 (±1.1) | 28.6 (±31.0) | 13.4 (±14.9) | 5.8 (±7.1) |
| **East grandstand** | 2.5 (±2.3) | 1.5 (±1.2) | 0.3 (±0.7) | 35.8 (±34.6) | 16.8 (±17.0) | 7.4 (±8.1) |
| **All stands** | 3.5 (±2.9) | 1.9 (±1.5) | 0.7 (±1.0) | 25.5 (±27.8) | 11.8 (±13.5) | 5.3 (±6.4) |

### Effects the event on epidemic spread of SARS-CoV-2 (simulation model)

In order to assess the potential effects of transmissions occurring during indoor events, we developed a dedicated individual based model. We investigated effects of MGE under conditions of epidemics controlled through overall reduction of contacts in the society and contact tracing of individuals with positive test results mimicking the current situation in Germany (reproduction number ∼1). In addition, we allowed for introduction of new cases (for example by persons who came back from regions with a higher incidence or due to super-spreading events) and in consequence, that number of new cases (incidence) is independent from the local transmission dynamics (represented by the reproduction number). In line with findings from serological studies, we assumed that current seroprevalence is negligible and most persons are susceptible to infection. We extrapolated the experimental data to reflect multiple events taking place in the city of Leipzig, assuming either 100000 or 200000 persons taking part in comparable MGEs per month (the latter estimate corresponded to the pre-pandemic state).

Whether transmissions take place in MGEs depends on how many infectious persons attend the event, how many contacts they have (including those related to spread of aerosols) and whether transmission is reduced, for example, due to masks. Excluding all individuals, either tested as positive, quarantined due to follow-up of other identified cases, or not tested but symptomatic, only the share of asymptomatic or pre-symptomatic persons, and those with symptoms not clear enough to be identified, will attend the event. For incidence of 50/100000 per week of positive tested cases, between 10 and 40 infectious persons on average will attend any event, assuming the total number of persons taking part in MGE is 100000 to 200000 per month (Fig. 3). Pre-pandemic contact numbers with an assumed reproduction number of around 3 result in transmission probability of about 7% per contact of 15+ minutes duration (based on German POLYMOD data (*19*). Consistently low are the resulting numbers of persons who would acquire the infection during the events (Fig. 4). For scenarios 2 and 3, particularly with the use of masks, the expected number of infections occurring in the events are below 10 per month. Consequently, the effect of the events on the total number of positive tested cases or quarantined persons when compared to the situation when no events take place is low (Fig. 5a). Overall, 2.3%, 1.1%, and 0.4% of the observed incidence would be attributed to MGEs for scenarios 1, 2 and 3 with the use of masks for the incidence of 100/100 000 per week and 100 000 persons in events per month.

**Fig. 3:**
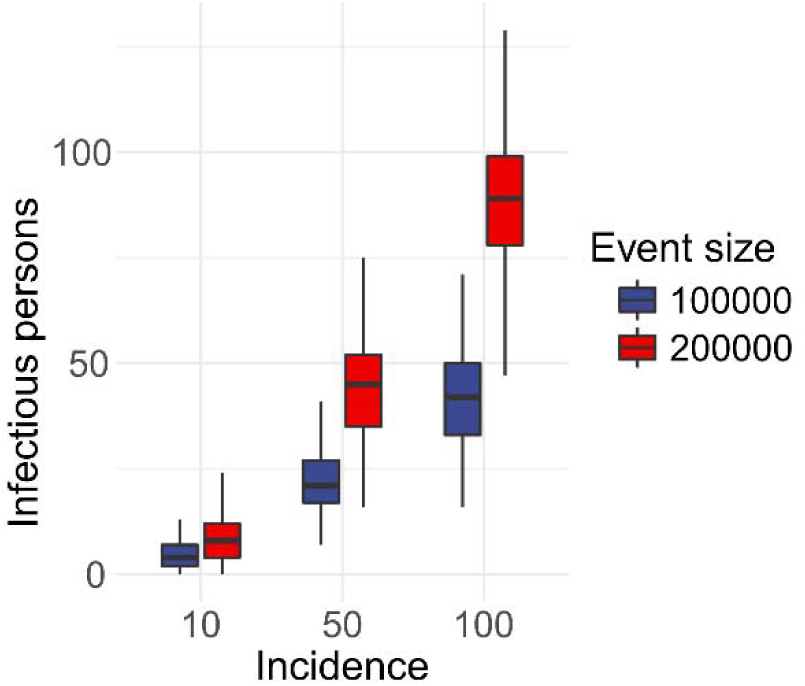
Expected number of infectious persons in MGE by total number of people attending events per month (event size) and incidence.

**Fig. 4:**
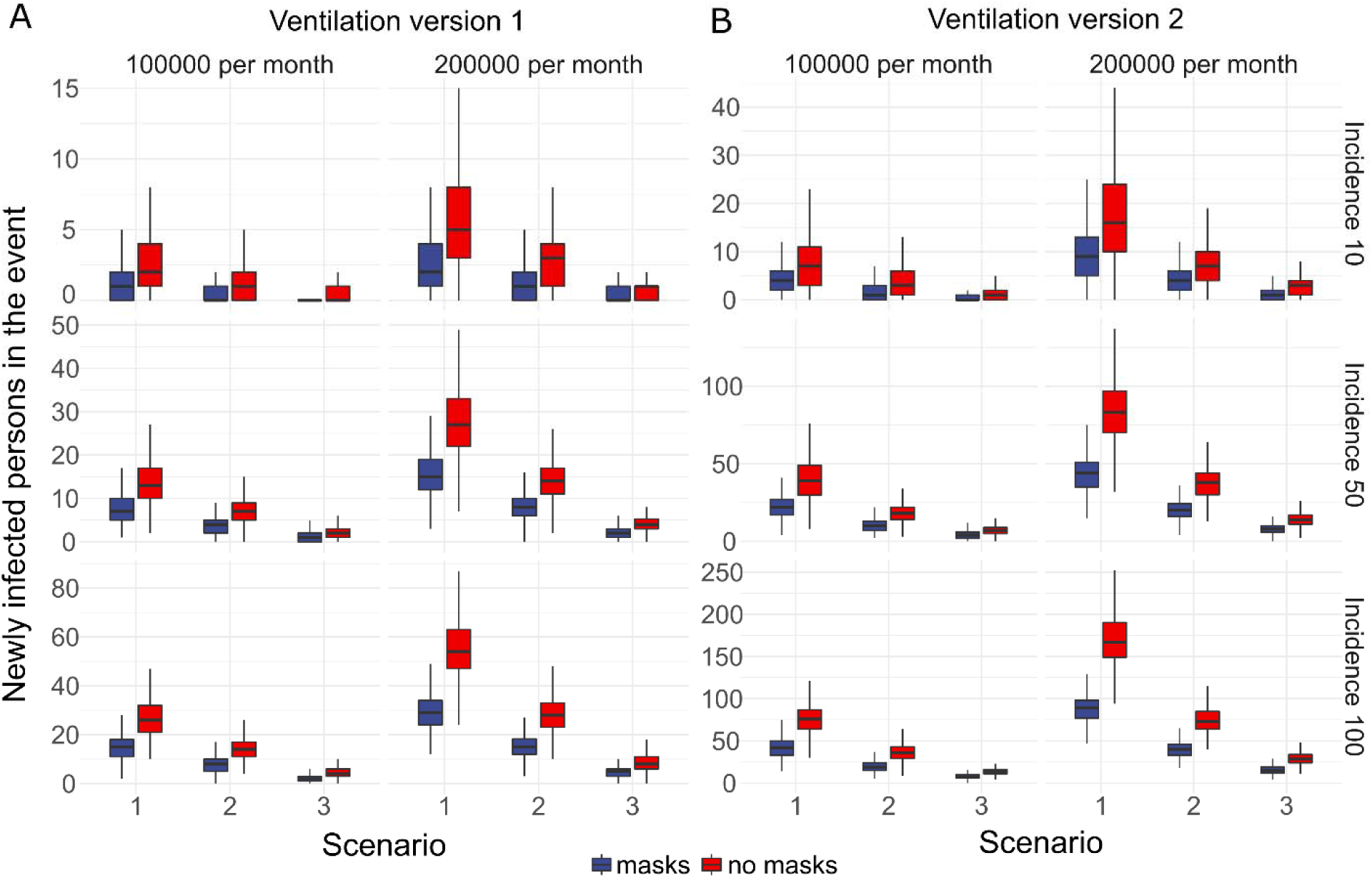
Number of persons who acquired the infection during MGE by scenario, participants in MGE per month, and comparison of masks vs. no masks for A) ventilation version 1, B) ventilation version 2

**Fig. 5:**
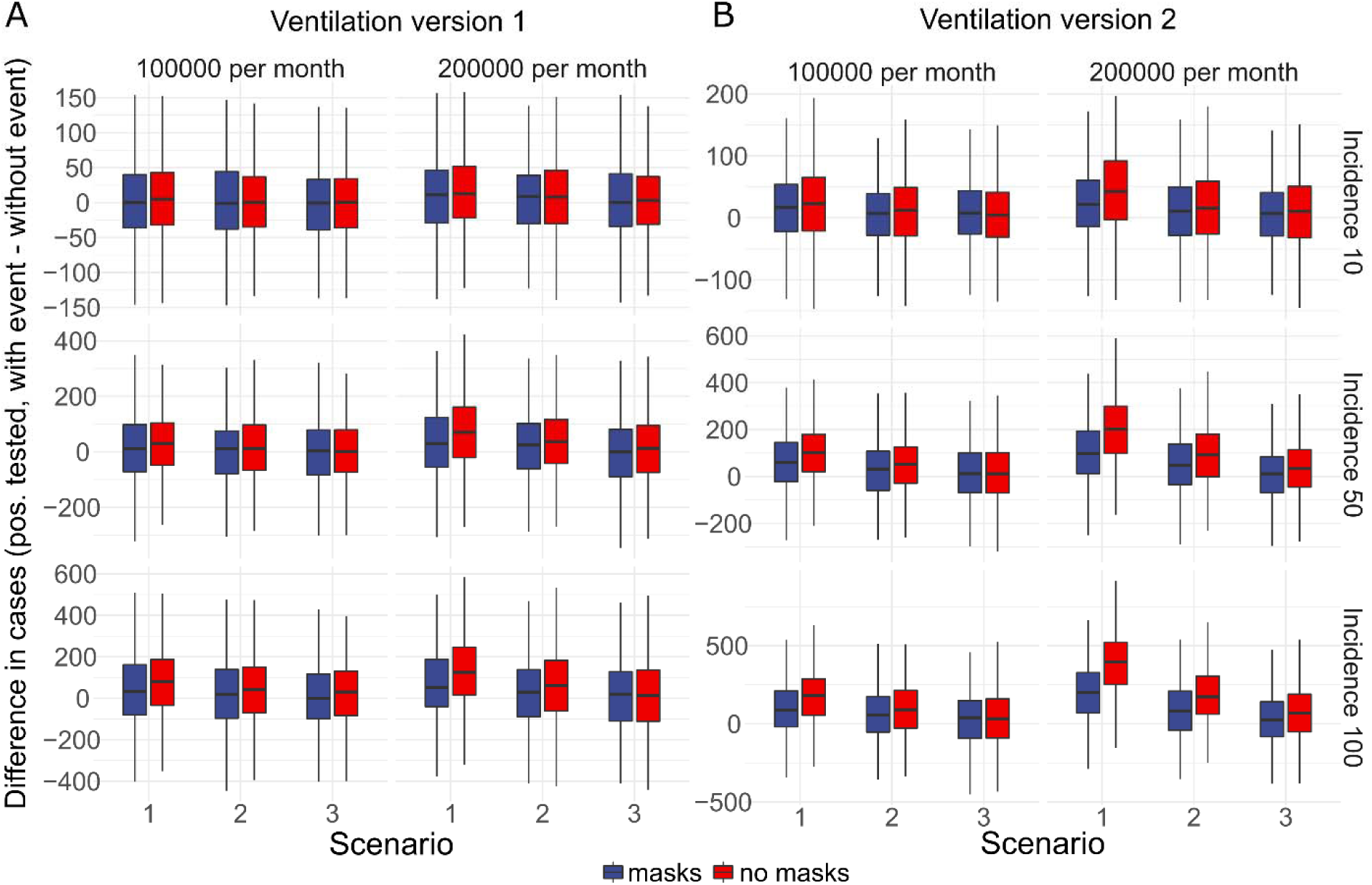
Comparison of the excess number of cases tested positive by scenario, participants in MGE per month, and comparison of masks vs. no masks for A) ventilation version 1 and B) ventilation version 2.

These results apply to the current ventilation in the arena (VV 1). However, when poor ventilation is assumed for all events taking place (VV 2), substantial additional burden of disease results (Fig. 5b). In the most negative scenario of those studied (200 000 persons in events per month, no masks in the event, no distancing – scenario 1, poor ventilation), the attributable proportion would increase to 23%. Further outcomes of the epidemiological simulation model are presented in Table S3.

### Acceptability of the hygiene concept (post-experiment survey)

Hygiene concept had strong effects on the number of contacts and potential transmissions. Therefore, we studied the perception of participants. A total of 960 participants completed the questionnaire (79% of all participants). Of those, 88% could picture attending an event or concert under the conditions of scenario 2 and 82% under the conditions of scenario 3. The majority of respondents (89%) felt that wearing N95-masks was unproblematic, sometimes a little restrictive, but they could get used to it quickly (Fig. S2). If it were necessary to wear normal mouth-nose protection or a N95 mask for a concert, 90% and 78% of the participants would do so.

## Discussion

Our results confirm the conventional wisdom that during MGEs, even without precautions, not every attendant has contact with all others. We also show that in scenarios with physical distancing, the resulting contact numbers are rather low and the effective risk depends primarily on the adequacy of the ventilation. Thus, under hygiene protocols and good ventilation, even a substantial number of indoor MGEs has only minimal effects on the overall number of infections in the population. However, poor ventilation systems can lead to a considerably higher rate of aerosol expositions and can thereby result in a high number of infections.

Compared to all contacts of the total population of inhabitants, MGEs contribute a small proportion of contacts taking place on any given day. In a setting of controlled epidemics (reproduction number at 1), additional contacts/infections can move the reproduction number of the epidemics above the threshold of 1. However, considering the small contribution of the events, it is unlikely that they will be the single cause for crossing the threshold. While in our simulations the difference with and without events is on average close to zero, in some cases, the numbers of new infections can be substantial. In an unfavourable case this may result in the impression that many infections were caused by the events. Apart from single “unfortunate” MGEs, which by chance resulted in outbreaks, MGEs without any precautions can have substantial contribution to epidemic spread. On the other side, MGEs under precautions contribute only a small fraction of new cases and this fraction would be maintained even if overall epidemic grows with R above 1. Furthermore, some of the contacts during MGEs might not be truly additional as the usual contact persons may be attending the event together.

While poor ventilation can substantially increase the number of transmissions, we expect that using masks and particularly N95 masks reduce the risk. In super-spreading events described in the literature, masks were not used (*20*). In the meantime, effects of masks on the reduction of transmission are meanwhile generally accepted (*21, 22*). Aerosols are of special concern during halves when visitors stay in their seats and exposition time can accumulate. Therefore, wearing masks for most of the time while sitting is mandatory to maximize the protective effect of masking.

With respect to contacts, particularly the “entry”, “half time” and “exit” phases are important, but particularly for the short time contacts, but in scenario 1 also for those of critical duration. Thus, hygiene concepts must address organizational aspects to keep contact times low in these periods (e.g. additional entrances, restricting eating and drinking to the seats, etc.).

In scenario 1, without any spacing between seats, the number of contacts (>15min) increased continuously over time, while the number of contacts stayed almost constant in scenarios 2 and 3. Thus, the use of seating plans including spacing rules (at least “checkerboard pattern seating”) are important to reduce contacts.

Two other potential measures are the use of tests before or dedicated CTDs during the event. Testing before an event adds additional security, but is very time and resource consuming. In addition, testing thousands of people within a few hours will be huge organizational challenge. In view of the overall low number of contacts with sufficient exposure and potentially much larger effect of aerosols which cannot be traced to individual persons, applying additional contact tracing of participants (when contact tracing devices are deployed during the event) also does not appear beneficial in the current setting. However, this might be beneficial during other kinds of mass gatherings.

Our results apply to MGEs with a sitting order and a high adherence to the implemented hygiene concept. Hygiene stewards rarely had to intervene in our experiment. This might be a consequence of the highly disciplined participants in our study. However, enforcing a hygiene concept in routine practice is crucial for risk reduction and can be supported by hygiene stewards.

Furthermore, large scale events (like soccer games) and standing concerts (e.g. rock concerts) might be different to the MGE we simulated with respect to the number of contacts and the probability of transmission (*21*). In the first, larger crowds will stand closer during the entry because of space restrictions and may gather additional contacts on the way to the event. In the latter, visitors have a high proximity to one another and do not stay on fixed positions. Therefore, the number of contacts can increase over time.

There are several limitations of our study. First, we did not reach our intended goal of 4000 participants of the event. We addressed this by space restrictions, but it is still possible that the density of contacts was reduced. Second, defining a threshold for a relevant aerosol exposition is quite difficult, as there are many unknown facts about airborne transmission of COVID-19 (e.g. the minimal infectious dose, the viral load of aerosols, etc.). However, since we used the same threshold for all calculations, results are consistent and comparable. Third, while we used a detailed model to simulate transmission, additional structures in the population can affect the results. For example, if only a small group will participate in all the events and transmit infections acquired in one event to another, this would results in higher impact of MGEs.

In conclusion, we found visitors of a seated concert in a good ventilated arena to have a high number of short contacts and a low number of long lasting contacts. Already moderate restrictive hygiene concepts (i.e. scenario 2) provided a substantial reduction of infections risk. Wearing masks during the concert was highly accepted by most participants and can provide further risk reduction. When hygiene concepts are applied and conditions of good ventilation are met, MGEs appear to contribute little to epidemic spread of COVID-19. Lack of hygiene concept and inadequate ventilation can increase the number of subjects at risk substantially.

## Supporting information

Vido S2

## Data Availability

Most of the quantitative results are provided in the supplemental tables. Distributions and individual level data in anonymized form can be obtained upon request. Code of the simulation model is submitted as supplementary material.

## Acknowledgments

First, we especially thank Karsten Guenther, managing director of SC DHfK Handball, Leipzig and Philipp Franke and Matthias Koelmel (directors of the Quarterback Immobilien Arena) who gave the impulse for setting up this study and passionately supported the experiment, provided the Arena and helped to recruit participants. Special thank goes to Frank Zimmermann (ZBP Zimmermann und Becker GmbH, Leipzig, Germany) who meticulously set up the CFD model and analysis. In addition, he kindly brought his long lasting expertise to the discussion of the results. We thank the musician Tim Bendzko who performed the concert. We also thank the governments of the Federal states of Saxony and Saxony-Anhalt who agreed to fund the project and the required tests. We thank all the helpers in the project, who prepared and organized the full day event. Finally, we would like to thank all participants of the experiment who donated their full day for this project.

## Funding

Ministry of economics, science and digitalization, federal state of Saxony-Anhalt and Ministry of science, culture and tourism, federal state of Saxony.

None of the involved parties had any influence on the authors with respect to the study results and the decision to publish those.

## Author contributions

**Stefan Moritz:** Conceptualization, Methodology, Formal analysis, Investigation, Resources, Writing - Original Draft, Writing - Review & Editing, Supervision, Project administration, Funding acquisition. **Cornelia Gottschick:** Conceptualization, Methodology, Investigation, Resources, Writing - Original Draft, Writing - Review & Editing, Visualization. **Johannes Horn:** Methodology, Software, Formal analysis, Investigation, Resources, Data Curation, Visualization. **Mario Popp:** Conceptualization, Methodology, Formal analysis, Investigation, Resources, Data Curation, Writing - Original Draft. **Susan Langer:** Investigation, Resources, Writing - Original Draft. **Bianca Klee:** Investigation, Resources, Writing - Original Draft. **Angelika Ihling:** Conceptualization, Resources. **Oliver Purschke:** Software, Formal analysis, Data Curation. **Michael Gekle:** Investigation, Supervision, Funding acquisition. **Rafael Mikolajczyk:** Conceptualization, Methodology, Investigation, Writing - Review & Editing, Supervision, Project administration.

## Supplementary Materials

Materials and Methods

Figures S1-S4

Tables S1-S9

Movies S1-S2

References (24-42)

