## Supplemental material for "The Risk of Indoor Sports and Culture Events for the Transmission of COVID-19 (Restart-19)"

**This PDF file includes:**

Materials and Methods

Figs. S1 to S4

Tables S1 to S2 and S4 to S9

**Other Supplementary Materials for this manuscript include the following:**

Movies S1 to S2

Table S3 as excel file

**Materials and Methods**

***General study design***

The “Risk prediction of indoor sports and culture events for the transmission of Covid-19” (RESTART-19) study was initiated in order to provide data on contacts and aerosol exposition at indoor mass gathering events (MGEs). The study had three components:

1) Experiment: In order to determine the number of contacts during a MGE, we conducted a pop concert under experimental conditions and provided all participants with a contact tracing device (CTD). The concert was conducted in three scenarios with different hygiene concepts.
2) Aerosol Distribution: For the assessment of aerosol exposition, a computer simulation of air flows was simulated.
3) Epidemiological Simulation: We integrated the results of contact tracing and aerosol distribution in an individual based model and simulated the effects on subsequent burden of infections.

The study protocol was submitted to the German clinical trial register (DRKS 00022790). (www.drks.de)

***1: Experimental concert simulation and contact measurement***

The event took place on August 22^nd^, 2020 in an indoor arena (Quarterback Immobilien Arena, QIA) in the city of Leipzig (Germany).

*Recruitment procedure and participants*

Individuals aged between 18 and 50 years were invited through extensive media campaign to register voluntarily and free of charge via the study webpage ([www.restart19.de](http://www.restart19.de)) where comprehensive information about the event, its objectives, and risks were provided. Participants did not receive any kind of allowance, but food and drinks were free throughout the day. A priori exclusion criteria were self-reported obesity (Body-Mass-Index >30), chronic diseases, and conditions affecting lungs, liver or kidneys, cardiovascular diseases, cancer, immune suppression, the intake of immunosuppressants, or pregnancy. We planned to include 4000 participants corresponding to half of the arena’s capacity and reflecting the mean event size for sports and culture events of the year 2019 at this location (4200 participants).

From July 17^th^ to August 21^st^, 2020 a total of 2825 participants registered for the study. 601 participants actively withdraw their consent and 212 participants did not confirm their registration. Thus, 2023 participants received the SARS-CoV-2 screening test set (see hygiene concept below). Of these, 1407 samples arrived on time in the laboratory for analysis and information on negative test was reported back to the participants. Only those with negative test result were asked to participate. One participant was tested positive and therefore excluded. In total, 1212 persons took part in the experiment.

*The event*

On the study day, all participants arrived between 8:00 and 10:00 a.m. for check-in. During check-in, participants were registered, identities confirmed and N95 masks, hand sanitizers, and contact tracing devices handed out for each person. Three tickets for three different scenarios were issued per person, containing information on timing, entrances, and seating.

We simulated three different scenarios in order to analyse the impact of different hygienic measures on the transmission of SARS-CoV-2. Each scenario followed the same schedule: entry (60 min), 1^st^ half (20 min), half time (20 min), 2^nd^ half (20 min), exit (15 min). During the halves, the German singer/songwriter Tim Bendzko performed a life pop concert. Scenarios differed with respect to hygiene measures such as number of entrances/exits, distance between seats, and restricted mixing of participants by dividing the arena into quadrants. Scenario 1 was designed to reflect a pre-pandemic state where participants entered and exited the arena through two main entrances without any restrictions and were seated without free seats in-between. Scenario 2 applied moderate hygiene measures: The arena was divided into four quadrants. Participants entered and exited the arena through the entrance/exit of the quadrant as indicated on their ticket (four entrances/exits) and were not allowed to change quadrants. A seating arrangement was implemented, where every second seat was occupied and the rows were shifted (checkerboard pattern). Scenario 3 reflects a stronger contact reduction, with pairwise seating of participants and implementation of a minimum distance of 1.5 m between occupied seats. In addition, the number of entrances/exits was increased to eight. The different scenarios are summarized in Table S4.

*The setting*

The Quarterback Immobilien Arena is an event location in the city of Leipzig with a seating capacity for up to 8228 people. Figure S3 shows an overall plan of the location. Visitors usually (i.e. before the pandemic) enter the hall via two main entrances (west and east side) opening into the foyer at the south end of the arena. From the foyer, they enter two long tunnels running parallel to the grandstands on each side of the hall. Visitors reach the grandstand via vomitories branching off the tunnel. In addition, the Arena has four emergency exits on each long side of the building, which were used in scenarios two and three to enter and exit the arena.

The total room volume of the Arena is 135000 m^3^. The ventilation system has a total capacity of 198 000 m^3^/h and uses 100% fresh air. The outlets under the grandstands have a capacity of 114 000 m^3^/h. In addition, there are jet nozzles above the heads of the spectators on the grandstand at the long sides, which blow air downstream to the inner space.

*Hygiene concept*

The Saxonian ministery of social affairs and cohesion (Sächsisches Staatsministerium für Soziales und Gesellschaftlichen Zusammenhalt) and the health authorities of the city of Leipzig approved the hygiene concept.

SARS-CoV-2 testing: One week before the event, all participants and staff members received a test set for SARS-CoV-2 including a swab and a tube containing stabilizing solution. The set included detailed instructions on self-sampling and on returning the test set. Participants were requested to take a throat swab within 48 hours before the event. The test sets could be dropped off at five different locations in Leipzig or Halle (Saale) or send via mail. All samples were analysed by the Institute of Virology of the University hospital in Leipzig. Test results were imported in the data bank the night before the event and participants received notification. Participants with positive or missing test results were informed by phone and were not allowed to enter the arena. The test was free of charge.

Exclusion criteria: On site exclusion criteria were no valid registration, no ID, positive or missing SARS-CoV-2 test, temperature above 37.5°C, self-reported symptoms of COVID-19 within the past 48 hours, contact to a COVID-19 patient or stay in risk area (according to Robert-Koch-Institute (RKI) of August, 22th 2020) within the last 14 days.

Personal protective equipment: During study-check-in, each participant received a N95 mask, a sanitizer bottle containing 85.5% Ethanol V/V as well as an ultra wide band-tracing device. The N95 mask had to be worn from entering to leaving the arena as well as in queues at the entrance, exits and simulated indoor catering stands.

Catering: Catering service took only place outside the arena where participants were allowed to remove their masks when a distance of 1.5 m was possible to keep. The catering was free of charge in order to reduce the waiting time for participants and to avoid participants leaving the area. Participants received on request water bottles inside the arena and were allowed to drink indoors given an appropriate distance to other people. During the half times in between the halves, indoor catering service was simulated in a way that people received vouchers to use outside.

Distance and hygiene stewards: Except for the first scenario, all participants were asked to keep a distance of 1.5 m. To ensure that all participants follow the hygiene concept, 40 hygiene stewards were present inside the arena. Participants repeatedly not adhering to the hygiene practice advices could be asked to leave the arena (but this was not necessary).

Briefing of participants and staff: All participants received comprehensive information regarding the hygiene concept upon registration. On the day of the event, participants received an information sheet with the hygiene rules and got instructions on proper use of N95 masks and hand sanitizers by the check-in staff. Furthermore, participants received verbal instructions at the beginning of the event. Staff received detailed training on hygiene concepts.

Contact tracing: During the registration process, participants provided full contact details. Participants agreed that their contact tracing devices (CTD) can be used to identify those at risk. In case of a SARS-CoV-2 infection after the event, affected participants would have been contacted. We are not aware of any cases who were infectious during the event. All personal informations were deleted six weeks after the event. The resulting data are anonymized.

Corona warn app: The use of the Corona-Warn-App of the German federal government was recommended, but not required for participation.

*Measurement of contacts in physical proximity*

All participants received a personalized contact tracing device (CTD) and were instructed to wear it around the neck during the event. The ICDWpro quad 164643 (In-Circuit, Dresden, Germany) tags were used to measure the distance between two participants and the time spent at this distance. These tags combine both Bluetooth low energy and ultra-wide band radio technology reaching an accuracy of +/- 20 cm. The CTD could either send or receive signals at any given time point so that there was an exchange of signals between the CTDs of all participants. The firmware and logging were customized according to the following protocol: a time stamp for the beginning of a contact was recorded if one of the following combinations of distance and time were met: <50 cm for at least 3 seconds, <100 cm for at least 6 seconds or <150 cm for at least 10 seconds. When leaving these thresholds for more than 2s a time stamp was logged. When the contact was broken, a new contact could be recorded after 10 sec reset time.

Due to the high number of tracers sending data simultaneously, a low broadcast intensity and small changes in distance movements of participants, the signals were often interrupted. Additionally, a CTD could only receive or send signals but not do both simultaneously. Thus, for a pair of sensors, proximity was recorded partly on the one and partly on the other. First, contacts from all devices were combined. Second, gaps were filled in between the first and last contact within a phase of the scenario (for example during halt time). Given this specification of the sensors, we were only able to use the information on the largest distance, i.e. 1.5 m (likely corresponding to a physical distance of 1.3 m when the sensors faced each other and less when signals were partly obscured by body parts). For a more realistic assessment of contacts, we scaled up the halves to 45 minutes each. In such way, also contacts with persons moving in and out of the radius of 1.5 during the sitting period could accumulate and cross the threshold of 15 minutes. We studied the total number of contacts lasting >10 sec (we included in this category contacts of >3 sec for the distance of 50 cm and >6 sec for the distance of 1 m) and 5 and 15 minutes. A critical contact was defined as lasting longer than 15 min within a distance of 1.5 m in line with contact definition by the Robert-Koch-Institute. For the overall number of contacts, the 15 minutes could accumulate through the full event.

*Acceptability questionnaire*

All participants of the experiment were asked via email to complete an online survey two weeks after the event. The questionnaire contained 10 questions regarding perception and opinion of the feasibility of such an event. The focus of interest was on wearing of masks and personal risk perception in the different scenarios.

**2: Aerosol distribution**

*Aerosol model*

Aerosol distribution within the arena was simulated using a computational fluid dynamics model. The fluid dynamics calculations were conducted with PHOENICS (Version 2020, CHAM, London, United Kingdom). For aerosol distribution the FLAIR module and for particle tracking the GENTRA-Tracking module were used. Validation of the used algorithm and further details are published on [www.cham.co.uk](http://www.cham.co.uk). The Quarterback Immobilien Arena was exactly transferred into a 3D-Modell with a 1:1 scale including all built-on components, the complete ventilation system, grandstands and seats. Virtual spectators were seated within the arena to simulate the aerosol emission and exposure. Since we expected very long calculation times (several weeks) for the whole simulation, we decided to model scenario 2 and to interpolate the other scenarios from this data. We installed 4000 virtual spectators within the model with a seating arrangement following a checkerboard pattern (i.e. every second chair remains free). 24 infectious persons were seated in 12 (out of 32) blocks. The detailed distribution of the infectious persons can be seen in Figure S3. The breathing air of all virtual spectators consists of an ideal gas and 20 litres CO_2_ per hour. In addition, infectious spectators emit aerosols of different sizes (0.5, 5 and 10 µm) into the room while breathing. Emission rate and particle size were adjusted to singing people (*23*). The physical density of respiratory air in g/ml was determined according to Hartmann et al. and Stadnytskyi et al. (*24*, *25*). For breathing volume and the corresponding proportion of CO_2_, we used a slightly increased value of 12 l/min (breathing air) and 20 l/h CO_2_ per person as we expected some excitement during singing, shouting or cheering. The detailed parameters and used equations for the model are summarized in Tables S8 and S9. To quantify the virus exposition of the spectators, each dummy was equipped with a virtual mouth which allows him/her to inhale air. Aerosol exposition was measured within the model directly at the mouth opening. To evaluate the dynamics and flow of the aerosol distribution within the arena, their aerosol movement was calculated via particle tracking and the results were presented through visualization and cumulated numbers. The numeric results on how the expired aerosols of the infectious participants spread in the arena were transferred into spreadsheets with the seating arrangement within the arena. From these spreadsheets, the number of affected persons and their level of exposure were obtained. We calculated an average value of persons exposed, in addition to those who would be captured by contact measurement with CTD. For visualization of the aerosol distribution, values below the critical threshold of 1.75 x 10-3 ng/s aerosol exposition (Fig. S1) were colored in green, whereas all values above are red. Brightness of the red color corresponds to the relative size of the mass flow.

*Variants of ventilation*

We ran the ventilation model in two different variants representing different ventilation systems. In the first ventilation variant, we modelled the actual ventilation system of the arena. Here, air reaches the arena via the outlets under the lateral grandstands and the jet nozzles as described above. For air suction, two towers are installed in each corner of the hall. The air supply was 198,000 m^3^/h corresponding to an air exchange rate of 1.46/h. In a second variant, we tried to optimize the ventilation system within the arena by virtually installing two long perforated suction tubes under the rooftop along the whole length of the arena. In addition, to avoid turbulences, the jet nozzles and the suction towers were switched off. In consequence, the air supply was reduced to 115,000 m³/h, corresponding to an air exchange rate of 0.85/h. Our goal was to improve layer ventilation and to reduce turbulences introduced by the nozzles, but as a side effect, the exchange rate was reduced.

*Defining participants with increased aerosol exposition*

The critical threshold for an aerosol exposition leading to an infection is not yet known. Several studies addressed this issue, but all of them have limitations, because many characteristics of SARS-CoV-2 (e.g. minimal infectious dose, virus concentrations in aerosols, etc.) are not yet known (*16*). Therefore, we used a pragmatic approach: A singing person emits about 1000 aerosol particles/s corresponding to 7.53x10^-8^ml of aerosols per second in our model (*23*). In contrast to that, a resting subject emits only a hundredth of particles (*16*). We assumed that the viral loads of aerosols are equal to sputum. In addition, the viral load is assumed to be high (10^9^ copies/ml) since infectious visitors are most likely to be presymptomatic or at the beginning of the symptomatic period and the viral load is known to peak at this time point (*26*, *27*). Thus, an infectious spectator emits 4x10^5^ virus copies (=7.53x10^-8^ml/s x 10^9^ virus copies/ml x 5400 s) when singing during a 90 min concert and 4x10^3^ virus copies (=4x10^5^/100) when resting. A threshold of 1% of the emission corresponds therefore to an exposure of 400 to 4000 virus particles per concert, which is a magnitude at which many assume the minimal infectious dose of Sars-CoV-2 (*28*).

**3: Epidemiological simulation**

*Natural history model*

The model was developed as an extended susceptible-exposed-infectious-recovered (SEIR) model (Fig. S4). Susceptible individuals are moving from the state of “exposed”, to “infectious presymptomatic” and “infectious” with a rate as indicated in Table S5. The disease of infectious individuals can either progress stepwise to the more severe stages “hospitalized”, “ICU admission”, “death” or “recovery” with an age-dependent probability (Table S6). A fraction of exposed remains asymptomatic and has a highly reduced infectiousness after the latent phase. Age-specific hospitalization rates were obtained from the federal state of Schleswig Holstein (*29*). Age-specific mortality was fitted to respective rates for Germany (*30*). Furthermore, we assumed the same per contact infection risk of 7% for all aerosol exposed individuals and direct contacts.

We simulated, in 600 runs, the combined contacts identified via contact tracing and aerosol distribution for all three scenarios with a baseline incidence of 10, 50, and 100 per 100,000 inhabitants/7 days, for an overall number of 100,000 and 200,000 participants in events per 30 days (corresponding to events with 3,300 and 6,700 participants per day, the latter number corresponding to the prepandemic state). We also studied scenarios including masks vs. no masks. For outcomes, a 30 day period was studied, which implicates that effectively the outcomes resulting from the events were counted over a shorter time period as some late outcomes might not develop within the studied time window.

*Contact network*

Model assumptions regarding daily contacts in the population were taken from the European contact study POLYMOD described in detail elsewhere (*19*). The age specific contact rates were applied to the population of the city of Leipzig (Table S6). For the model, we consider three types of contact settings: “household”, “school/work” and “other” (including the contact categories transport, leisure and other from the POLYMOD study (*19*), Table S7). We assume that for each person there is exactly one home place (household). For each person aged 0-19 years, there is exactly one place at day-care or school. Each school or day care class consists of 20 children (reflecting the average class size in Saxony), as well as one teacher 20 to 64 years old. For each person between age of 20 and 69 years (apart from teachers), there is one work place. We included four different workplace sizes according to the categories reported by the Federal Statistical Office in Germany (*31*). The selection of the working place for a person is random but depends on age (based on additional analyses of POLYMOD data). For the numbers of contacts for each person, we only considered contacts longer than 15 minutes, which is in line with the high-risk contact definition from the RKI.

*Epidemic control measures*

Our model is based on the national guidelines of Germany regarding the SARS-CoV-2 testing strategy, i.e. testing due to symptoms or as a contact person of a known case. We assume perfect tests with 100% sensitivity and specificity. In the model, there are two ways to detect infected individuals, either due to symptoms or due to contact tracing. A case with a severe course of COVID-19 is assumed to be tested, detected, and isolated within one day after symptom onset with an overall probability of 90%. Cases with mild symptoms are assumed to be tested within two days after symptom onset with an overall probability for testing of 50%. Asymptomatic individuals, who never develop symptoms, can only be detected by contact tracing. After a person is tested positive, the household members of this individual are requested to be tested within one day and stay in quarantine for 14 days. We did not account for non-compliance during the self-quarantine. We assumed a detection rate of 100% for all household members, thus adding new branches for contact tracing. In contrast, the testing rate is assumed to be 80% within the school or work network with a delay of 2 days for the test result. In case of the third contact network “other”, we assumed that only 50% of contacts were identified and tested with a delay of four days. Infected persons are detected through contact tracing, if the individual passed the latent phase, including pre-symptomatic and completely asymptomatic persons. Contact tracing is triggered by the new detection of any infected person due to symptoms or past contact tracing. Recursive contact tracing can therefore result in the detection of whole infection chains. Any detected person is isolated for 14 days (including hospital stay if necessary). In this model, secondary contacts were not quarantined pre-emptively, which is in line with the national test strategy of Germany, but could be identified once the primary contact became a confirmed case.

*Reproduction number*

The POLYMOD contact matrix corresponds to the pre-pandemic state. We calibrated the average per contact transmission probability to obtain a reproduction number for the epidemics of about 3 (while assuming the susceptible fraction at 100% - i.e. conditions at the beginning of the pandemic) and subsequently reduced the contacts in all settings uniformly by 50% and applied the epidemic control measures finally arriving at R around 1. The POLYMOD contact matrix ignores potential transmission of respiratory pathogens due to aerosols, which increases the probability of acquiring infection in a contact for a given reproduction number.

*Demographic background*

The study was conducted in Leipzig, which is a large city in the Federal State of Saxony in eastern Germany with about 601.083 inhabitants (*32*). Detailed demographics for the model are based upon Leipzig (*32*). Since the duration of the simulated epidemic is less than a year, we do not consider changes in the population due to births, deaths migration or aging.

***Statistical Analysis***

For statistical analyses, the mean and the standard deviation around the mean, the range and interquartile range (IQR) were calculated.

***Ethics Statement***

The Ethics Committee of the Martin-Luther-University (Halle, Germany) approved the RESTART-19 study. The responsible authorities (Saechsisches Staatsministerium fuer Soziales und gesellschaftlichen Zusammenhalt) and the Public Health Authority of the city of Leipzig permitted the study according to the submitted hygiene and safety concept.

**Figures and Tables**

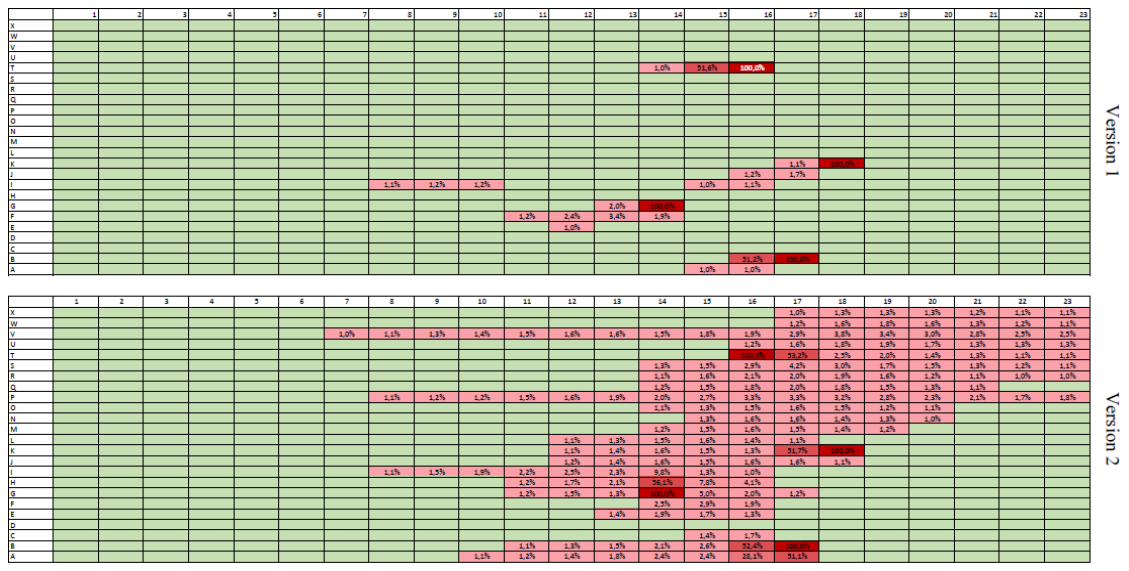

**Fig. S1:** Two Seating blocks within the arena with amount of aerosol exposition resulting from infectious individuals (indicated in dark red with black frame). People on green colored seats will receive <1% of the emitted aerosol amount. Red colored seats get more than 1%. Current ventilation version 1 (upper part), simulated ventilation version 2 (lower part).

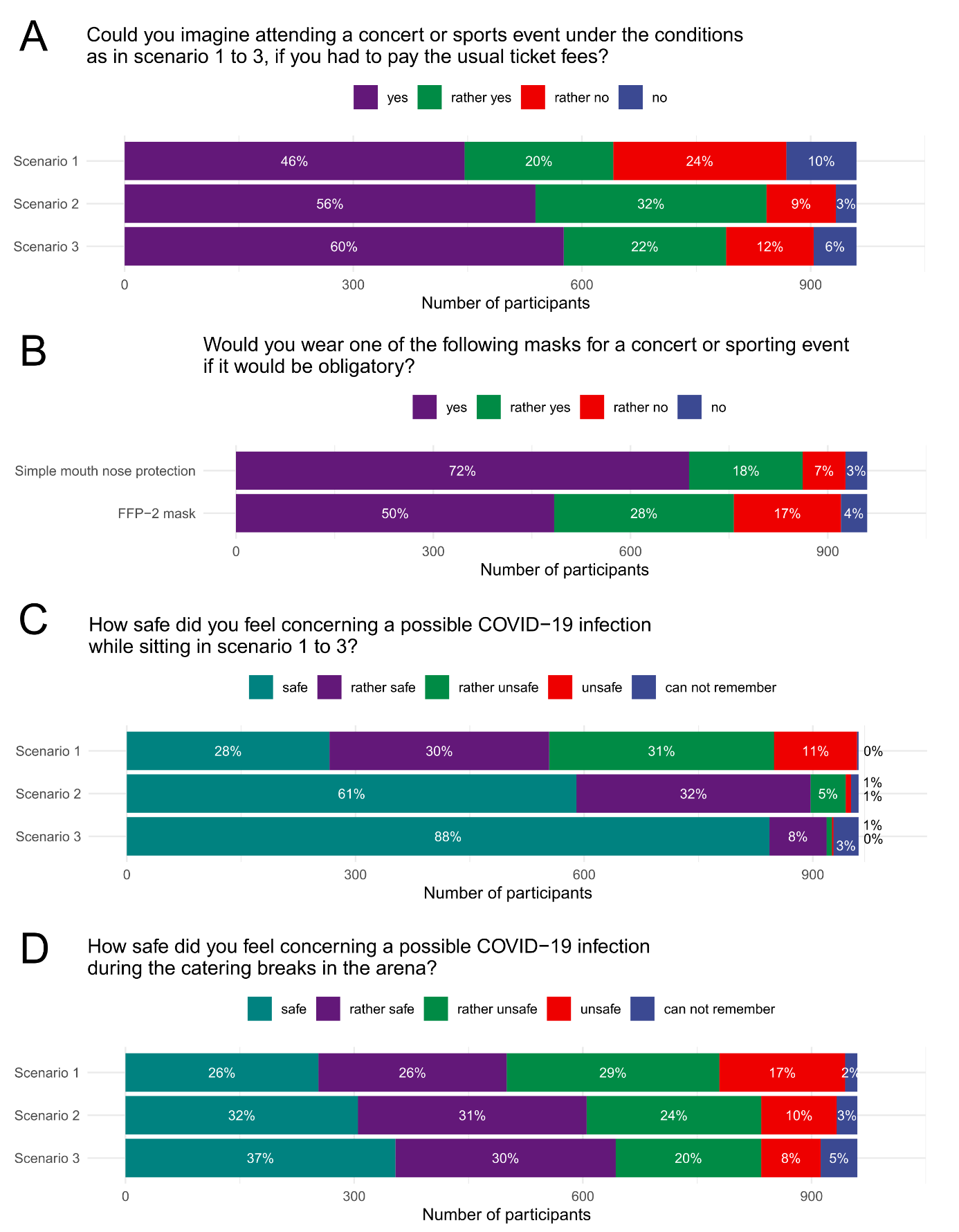

**Fig. S2:** Results of the survey.

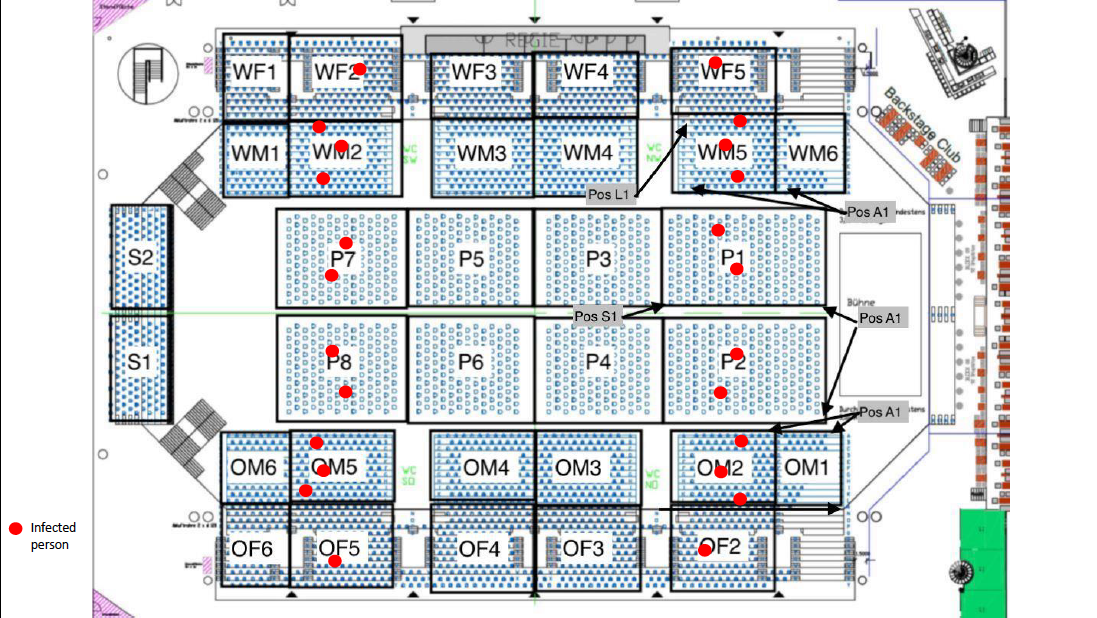

**Fig. S3:** Distribution of infectious persons within the arena for the aerosol simulation. Red dots indicate infectious persons. WF: upper west grandstand; WM: lower west grandstand; OF: upper east grandstand; OM: lower east grandstand; P: floor

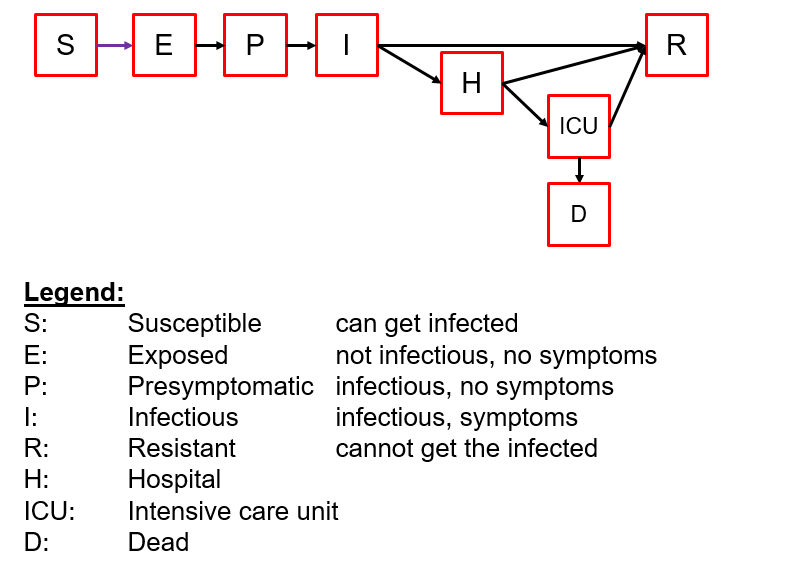

**Fig. S4:** Extended SEIR model.

**Table S1.** Socio-demographic characteristics of the participants of the live event (n=1212).

| Variables | N (%) |
| --- | --- |
| Age  18-25  26-30  31-35  36-40  41-45  46-50 | 355 (29.3)  169 (13.9)  248 (20.5)  186 (15.3)  143 (11.8)  111 (9.2) |
| Sex  Male  Female  Diverse | 443 (36.6)  767 (63.3)  2 (0.2) |
| Area of residence  Leipzig  Saxony (outside Leipzig)  Outside Saxony | 471 (38.9)  404 (33.3)  337 (27.8) |

**Table S2.** Mean number of measured contacts using mobile contact tracing devices. Total contacts longer than 10s, 5min and 15 minutes are shown as well as contacts stratified by setting (entry, 1^st^ half, half time, 2^nd^ half, exit) within all scenarios (1, 2, and 3) and cumulatively. SD = standard deviation, Sc = scenario.

| Time  cut-off | Sc | Mean number of measured contacts (± SD) | | | | | |
| --- | --- | --- | --- | --- | --- | --- | --- |
|  |  | Total | Entry | 1^st^ half | half time | 2^nd^ half | Exit |
| ≥ 10s | 1 | 63.9 (±17.1) | 30.8 (±12.1) | 7.0 (±2.8) | 24.5 (±9.2) | 6.4 (±2.7) | 15.3 (±6.7) |
|  | 2 | 36.4 (±12.0) | 13.5 (±5.3) | 3.7 (±1.5) | 14.7 (±6.7) | 4.3 (±1.8) | 14.9 (±7.6) |
|  | 3 | 18.0 (±7.2) | 6.1 (±3.5) | 1.3 (±0.6) | 8.3 (±5.1) | 1.8 (±1.5) | 6.1 (±3.6) |
| ≥ 5min | 1 | 14.1 (±5.2) | 8.7 (±4.1) | 5.3 (±2.3) | 3.1 (±2.4) | 4.4 (±2.0) | 6.4 (±1.4) |
|  | 2 | 6.1 (±2.4) | 4.9 (±2.1) | 2.7 (±1.3) | 2.6 (±1.5) | 3.2 (±1.5) | 1.1 (±0.9) |
|  | 3 | 2.2 (±1.5) | 2.0 (±1.3) | 1.0 (±0.3) | 1.2 (±1.9) | 1.0 (±0.7) | 0.7 (±0.7) |
| ≥ 15min | 1 | 8.9 (±3.5) | 5.1 (±2.5) | 4.5 (±2.1) | 1.8 (±1.3) | 3.9 (±1.9) | 0 (±0) |
|  | 2 | 4.7 (±1.9) | 3.7 (±1.6) | 2.3 (±1.2) | 1.9 (±1.2) | 2.9 (±1.4) | 0 (±0) |
|  | 3 | 1.3 (±0.9) | 1.1 (±0.6) | 1.0 (±0.3) | 0.8 (±0.7) | 0.9 (±0.6) | 0 (±0) |
| ≥ 10s  cumulative | 1 |  | 30.9 | 32.8 | 52.1 | 54.1 | 63.9 |
|  | 2 |  | 13.5 | 13.9 | 24.7 | 25.3 | 36.4 |
|  | 3 |  | 6.1 | 6.3 | 13.3 | 14.1 | 18.0 |
| ≥ 5min  cumulative | 1 |  | 8.7 | 11.0 | 12.3 | 13.7 | 14.1 |
|  | 2 |  | 5.0 | 5.2 | 5.6 | 6.0 | 6.1 |
|  | 3 |  | 2.0 | 2.0 | 2.1 | 2.2 | 2.2 |
| ≥ 15min  cumulative | 1 |  | 5.1 | 7.3 | 7.5 | 8.9 | 8.9 |
|  | 2 |  | 3.7 | 4.0 | 4.2 | 4.7 | 4.7 |
|  | 3 |  | 1.1 | 1.1 | 1.3 | 1.3 | 1.3 |

**Table S3.** Epidemiological outcomes (submitted separately as excel file)

**Table S4.** Components of the hygiene concepts in the three different scenarios.

|  | Scenario 1 | Scenario 2 | Scenario 3 |
| --- | --- | --- | --- |
| Seating | No seats free | Every 2^nd^ seat free  “Checkboard pattern” | Pairwise,  1.5 m around free |
| Quadrants | No | Yes | Yes |
| Entrances | 2 | 4 | 8 |
| Catering | unrestricted | In Quadrants | In Quadrants |
| Toilets | unrestricted | Every 2^nd^ urinal closed | Every 2^nd^ urinal closed |

**Table S5.**Parameters for transmission model

E=exposed individuals (not infectious, no symptoms), P=pre-symptomatic individuals (infectious, no symptoms), I=infectious individuals (infectious, symptoms), H=hospital submission, ICU=submission to intensive care unit, D=death, R=resistant individuals.

| From | To | Assumed mean value in days (n; probability) | Details | Ref. |
| --- | --- | --- | --- | --- |
| E | P | 3 (4; 0.5) | latent period ranged from 2 to 4 days | (*33*, *34*) |
| P | I | 1.5 (2; 0.25) | duration of asymptomatic infectiousness ranged from 1 to 3 days | (*35*, *36*) |
| I | H | 4 (6; 0.5) | duration from having symptoms to being hospitalized was 4 days | (*37*) |
| H | ICU | 2 (2; 0.5) | duration from hospitalization to ICU ranged from 1 to 3.5 days | (*34*, *38*) |
| ICU | D | 8 (10; 0.7) | time from ICU to death ranged from 3.5 to 14 days | (*33*) |
| I | R | 6 (10; 0.5) | 5 to 10 incl. presymptomatic | (*39*) |
| H | R | 11.8 (12; 0.9) | duration from hospital admission to recovery ranged from 10 to 14 days (for non ICU-patients) | (*40*) |
| ICU | R | 12 (22; 0.5) | duration from ICU to recovery ranged from 8 to 17 days | (*33*, *40*) |

**Table S6.** Fractions progressing from one state to another divided into age groups and individuals modelled per age group.

| Age group | Fraction of no symptoms (E🡪R) | Fraction hospitalized (I🡪H) | Fraction in ICU per Hospitalized (H🡪ICU) | Lethality (ICU🡪D) | Number of individuals modelled (*32*) |
| --- | --- | --- | --- | --- | --- |
| 0-4  5-9  10-14  15-19  20-24  25-29  30-34  35-39  40-44  45-49  50-54  55-59  60-64  65-69  70-74  75-79  80-84  85-++ | 17% for all age groups | 10.0%  10.0%  10.0%  10.0%  10.0%  10.0%  12.0%  12.0%  15.0%  15.0%  21.0%  21.0%  27.0%  27.0%  39.0%  39.0%  53.0%  53.0% | 20.84%  9.32%  9.10%  6.77%  7.02%  14.18%  14.81%  8.89%  13.86%  17.24%  11.55%  19.13%  22.85%  38.93%  30.28%  45.10%  35.55%  28.22% | 6.60%  6.60%  5.0%  5.0%  10.0%  10.0%  25.0%  25.0%  25.0%  25.0%  62.73%  62.73%  59.09%  59.09%  80.0%  80.0%  80.0%  80.0% | 31497  27905  23801  24831  42208  48165  59903  49607  38700  33017  36880  37173  30703  30181  21741  29420  23872  17355 |
| Ref | (*41*) | Adjusted to fit to hospitalization rate in Schleswig-Holstein (*29*) | adjusted/multiplied to fit to the total number of ICU admissions in Germany (*42*) | Adjusted to fit to mortality rate in Germany (*30*) |  |

**Table S7.** Number of contacts per household size, setting and age group.

| Age group | Type of contact (*19*) | | | | | | | | | | | | | | | | |
| --- | --- | --- | --- | --- | --- | --- | --- | --- | --- | --- | --- | --- | --- | --- | --- | --- | --- |
|  | Household size of 1 | | | | Household size of 2 | | | Household size of 3 | | | Household size of 4 | | | | Household size of 5 | | |
|  | House-hold | School/ work | Other | House-hold | School/ work | Other | House-hold | School/ work | Other | House-hold | | School/ work | Other | House-hold | | School/ work | Other |
| 0-4  5-9  10-14  15-19  20-24  25-29  30-34  35-39  40-44  45-49  50-54  55-59  60-64  65-69  70-74  75-79  80-84  85-++ | 0.93  0.93  0.93  0.73  0.73  0.73  0.73  0.73  0.73  0.73  0.73  0.73  0.73  0.73  0.73  0.73  0.73  0.73 | 1.44  1.65  1.88  2.15  1.12  1.12  1.12  1.12  1.12  1.12  1.12  1.12  0.15  0.15  0.00  0.00  0.00  0.00 | 1.19  1.35  1.53  1.74  1.78  1.65  1.53  1.42  1.32  1.22  1.13  1.05  0.98  0.91  0.84  0.78  0.72  0.67 | 1.15  1.15  1.15  0.93  0.93  0.93  0.93  0.93  0.93  0.93  0.93  0.93  0.93  0.93  0.93  0.93  0.93  0.93 | 1.23  1.41  1.61  1.83  1.12  1.12  1.12  1.12  1.12  1.12  1.12  1.12  0.15  0.15  0.00  0.00  0.00  0.00 | 1.19  1.35  1.53  1.74  1.78  1.65  1.53  1.42  1.32  1.22  1.13  1.05  0.98  0.91  0.84  0.78  0.72  0.67 | 1.41  1.41  1.41  1.20  1.20  1.20  1.20  1.20  1.20  1.20  1.20  1.20  1.20  1.20  1.20  1.20  1.20  1.20 | 1.05  1.20  1.37  1.56  1.12  1.12  1.12  1.12  1.12  1.12  1.12  1.12  0.15  0.15  0.00  0.00  0.00  0.00 | 1.19  1.35  1.53  1.74  1.78  1.65  1.53  1.42  1.32  1.22  1.13  1.05  0.98  0.91  0.84  0.78  0.72  0.67 | 1.73  1.73  1.73  1.53  1.53  1.53  1.53  1.53  1.53  1.53  1.53  1.53  1.53  1.53  1.53  1.53  1.53  1.53 | | 0.90  1.02  1.17  1.34  1.12  1.12  1.12  1.12  1.12  1.12  1.12  1.12  0.15  0.15  0.00  0.00  0.00  0.00 | 1.19  1.35  1.53  1.74  1.78  1.65  1.53  1.42  1.32  1.22  1.13  1.05  0.98  0.91  0.84  0.78  0.72  0.67 | 2.12  2.12  2.12  1.96  1.96  1.96  1.96  1.96  1.96  1.96  1.96  1.96  1.96  1.96  1.96  1.96  1.96  1.96 | | 0.77  0.87  1.00  1.14  1.12  1.12  1.12  1.12  1.12  1.12  1.12  1.12  0.15  0.15  0.00  0.00  0.00  0.00 | 1.19  1.35  1.53  1.74  1.78  1.65  1.53  1.42  1.32  1.22  1.13  1.05  0.98  0.91  0.84  0.78  0.72  0.67 |

**Table S8.** Parameters and equations used for the CFD Model.

| **Parameters and equations** | **Details** |
| --- | --- |
| n | number of particles in the respiratory air per second |
| with n | number of aerosols |
| ρ_p_ = 1300 | density of the aerosol in kg/m^3^ |
| V_p_ = 4/3*π*(D/2)^3^ | particle volume in m^3^ |
| V = n*V_p_ | entire volume flow particle type |
| m = V* ρ_p_ | mass flow particle type |
| T = 31.5°C | breathing temperature |
| P1 = 100.13 | air pressure in kPa |
| rh = 0.5 | relative humidity 50% |
| V_CO2_ = 0.020 / 3600 | = 20 litre/h CO_2_ in m3/s |
| Lung Volume = 4.5*1.8 | = 12 breaths/minute = 8.1 litre/min |
| V_breath_ = 0.0081 / 60 | = 0.000135 m^3^/s |
| ρ_breath_ = ρ(AirH_2_O; T = T; R = rh; P = P1) = 1.135 | density of respiratory air in kg/m3 |
| m_breath_ = V_breath_*ρ_breath_ = 0.0001532 | mass flow of breath in kg/s |
| ρ_CO2_ = ρ(CO_2_; T = T; P = P1) = 1.74 | density of CO_2_ in kg/m3 |
| m_CO2_ = VCO_2_*ρCO_2_ | = 0.000009665 kg/s |
| m_sum_ = 5.27*10^-9^ | sum column 4, aerosols in kg/s |
| m_total_ = m_sum_ + m_breath_ + m_CO2_ = 0,0001629 | total mass flow in kg/s |
| Ai = m / m_total_ | amount of mass flow |
| A_breath_ = m_breath_ / m_total_ = 0,9406 | amount respiratory air |
| A_CO2_ = m_CO2_ / m_total_ = 0.05933 | amount CO_2_ |
| A_control_ = A_breath_ + A_CO2_ + 32.4*10^-6^ = 1.0 | sum (column 5) Ai = 32.4*10^-6^ |

**Table S9.** Particle distribution used for the aerosol model. D = diameter of the aerosols; n = number of particles in the respiratory air per second; V = particle volume; m = mass flow; A_i_ = amount of mass flow

| Run | *D* [m] | *n* [1/s] | *V* [m^3^/s] | *m* [kg/s] | *A_i_* |
| --- | --- | --- | --- | --- | --- |
| 1 | 5.000E-07 | 550 | 3.600E-17 | 4.680E-14 | 287E-12 |
| 2 | 5.000E-06 | 350 | 2.291E-14 | 2.978E-11 | 183E-09 |
| 3 | 1.000E-05 | 100 | 5.236E-14 | 6.807E-11 | 418E-09 |
| 4 | 3.000E-05 | 50 | 7.069E-13 | 9.189E-10 | 5.6E-06 |
| 5 | 5.000E-05 | 50 | 3.272E-12 | 4.254E-09 | 26E-06 |

**Movie S1:**

Particle Tracking VV1

**Movie S2:**

Particle Tracking VV2
